# Artificial Intelligence Performance in Testing Microfluidics for Point-of-Care

**DOI:** 10.1101/2024.06.24.24309435

**Authors:** Mert Tunca Doganay, Purbali Chakraborty, Sri Moukthika, Soujanya Jammalamadaka, Dheerendranath Battalapalli, Mohamed S. Draz

## Abstract

Artificial intelligence (AI) is revolutionizing medicine by automating tasks like image segmentation and pattern recognition. These AI approaches support seamless integration with existing platforms, enhancing diagnostics, treatment, and patient care. While recent advancements have demonstrated AI superiority in advancing microfluidics for point of care diagnostics, a gap remains in comparative evaluations of AI algorithms in testing microfluidics. We conducted a comparative evaluation of AI models specifically for the two-class classification problem of identifying the presence or absence of bubbles in microfluidic channels under various imaging conditions. Using a model microfluidic system with a single channel loaded with 3D transparent objects (bubbles), we challenged each of the tested machine learning (ML) (n = 6) and deep learning (DL) (n = 9) models across different background settings. Evaluation revealed that the Random Forest ML model achieved 95.52% sensitivity, 82.57% specificity, and 97% AUC, outperforming other ML algorithms. Among DL models suitable for mobile integration, DenseNet169 demonstrated superior performance, achieving 92.63% sensitivity, 92.22% specificity, and 92% AUC. Remarkably, DenseNet169 integration into a mobile POC system demonstrated exceptional accuracy (> 0.84) in testing microfluidics at under challenging imaging settings. Our study confirms the transformative potential of AI in healthcare, emphasizing its capacity to revolutionize precision medicine through accurate and accessible diagnostics. The integration of AI into healthcare systems holds promise for enhancing patient outcomes and streamlining healthcare delivery.

## INTRODUCTION

The convergence of artificial intelligence (AI) and healthcare has opened up a new era of possibilities, particularly in detection diagnostics and treatment. With AI algorithms continuously advancing, the integration of these approaches into healthcare systems holds immense promise for transforming traditional practices and addressing longstanding challenges in healthcare delivery[1–3]. Healthcare applications driven by sophisticated machine learning (ML) and deep learning (DL) algorithms stand at the forefront of modern healthcare innovation[4–6]. These algorithms empower machines to obtain insights from vast datasets, predict clinical outcomes, and assist healthcare providers in making informed decisions[6]. From medical imaging analysis to personalized treatment strategies, AI-driven approaches have demonstrated significant efficacy in improving diagnostic precision and ultimately enhancing patient outcomes[7–10].

Point-of-care diagnostics represent a cornerstone of modern healthcare, offering timely and accessible testing solutions, particularly in resource-limited settings[11–13]. The integration of AI into microfluidic systems presents a promising avenue for enhancing the accessibility and efficiency of POC testing[14, 15]. By harnessing advanced ML and DL algorithms, AI enhances the sensitivity, specificity, and multiplexing capabilities of microfluidic devices, enabling rapid and accurate detection of a wide range of diseases and biomarkers directly at the point of care[16–18]. An important approach where AI is utilized to enhance microfluidic systems is in facilitating its integration with other approaches and image processing[19–21]. ML and DL learning models excel at image classification and pattern recognition tasks and can support microfluidic devices to perform rapid and multiplex assays, allowing for comprehensive screening or testing using minimal resources[22–24]. This integration addresses critical gaps in healthcare access and empowers a new level of POC diagnostics, equipping frontline providers with actionable insights and revolutionizing the delivery of healthcare services.

Recent advancements have demonstrated superior performance in identifying disease biomarkers, detecting cancer[25], viruses[26], bacteria[27], and other pathogens[28], underscoring the robustness and clinical relevance of AI-integrated microfluidic platforms in modern healthcare settings. However, despite these advancements, there remains a gap in the comparative evaluations of different AI algorithms in testing microfluidics, and the optimal approach for maximizing their performance in this context remains unclear[29–34]. To address this gap, this study aims to critically analyze existing AI algorithms in testing microfluidics, focusing on their efficiency in enabling microfluidic platforms and their implications on diagnostic accuracy, resource utilization, and scalability. In the context of POC diagnostics, where real-world deployment considerations such as cost, power consumption, and scalability are paramount, the choice of algorithm can have significant implications. Such a comparative study is critical in gaining a comprehensive understanding of the strengths and weaknesses of different algorithms, informing algorithm selection, optimization, and deployment decisions across diverse domains and applications[35–38].

We employed a model microfluidic system, featuring a single microfluidic channel loaded with 3D transparent objects of bubbles. This model is designed to rigorously challenge the performance of commonly used AI models and provide insights into their effectiveness in real-world diagnostic scenarios. We integrated various ML and DL algorithms into our study, including CNNs like MobileNetV2, ResNet101V2, and DenseNet169, alongside commonly used ML models in healthcare applications such as Naive Bayes, logistic regression, KNN, SVM, and Random Forest[39–43]. Among the six evaluated ML algorithms, the Random Forest model performed best, achieving 95.52% sensitivity, 82.57% specificity, and 97% AUC. Similarly, among the nine DL models, DenseNet169 stood out, achieving 92.63% sensitivity, 92.22% specificity, and 92% AUC.

## RESULTS AND DISCUSSION

The integration of AI in medicine is driven by its remarkable ability to analyze and classify images and datasets. This computational capability of AI algorithms is foundational across diverse domains, prominently within diagnostics and medical testing, where AI-driven image analysis stands as a transformative force, providing rapid data processing and precise assessment devoid of infrastructure constraints or specialized human oversight[3, 44, 45]. This technological paradigm bears profound implications, particularly on POC diagnostics, through its role in facilitating the integration of microfluidics into POC applications[46]. By harnessing sophisticated ML and DL algorithms, AI streamlines the imaging and analysis of microfluidic devices, such as smartphone-captured assays, reducing the total testing cost and time, enhancing accuracy, and expanding utility[22, 47, 48]. This convergence of AI and microfluidics within POC holds immense potential to democratize healthcare access, particularly in underserved regions, by providing affordable, accurate, and accessible diagnostic solutions[14, 22, 49, 50].

In our study, we investigated the efficacy of AI algorithms, including both ML and DL, to facilitate the process of testing microfluidics within POC settings. We employed a microfluidic system comprising a single microfluidic channel to rigorously assess a set of 15 AI models recognized for data analysis and image classification across biomedical and diagnostic domains. Our experimental setup incorporated testing configurations featuring varying densities of bubbles. Bubbles as a readout was selected to probe the imaging and analytical performance of the examined algorithms. Despite bubbles being less prevalent than conventional color-based or fluorescence-based readouts, their inherent 3D transparency poses challenges, as they may be mistaken for non-targeted constituents within the sample matrix, microfluidic system or the testing environment and background. Colorimetric readouts, though linear and would allow comparatively easier workflow, fail to sufficiently encapsulate the intricacies necessary for discerning strengths and weaknesses of the tested algorithms. Meanwhile, fluorescence, although know to support high specificity and sensitivity testing, remains impractical for widespread POC adoption due to the need for bulky equipment and specialized setup to achieve the required sensitivity and specificity in most analyses.

Our set of AI algorithms included ML models, such as Naive Bayes, logistic regression, k-Nearest Neighbors (KNN), Support Vector Machines (SVM), and Random Forest, alongside DL CNNs such as MobileNetV2, ResNet101V2, and DenseNet169. By combining traditional ML algorithms with state-of-the-art CNN architectures, we created a diverse ensemble of models that can collectively leverage different aspects of the data. This ensemble approach is essential to enhance robustness and generalization performance, particularly in scenarios where the dataset may be limited or the target features are challenging to discern (i.e., bubbles). The incorporation of traditional ML algorithms stemmed from their robustness in handling various types of features, including those extracted from images, and their suitability for the often constrained datasets characteristic of microfluidic diagnostics at POC settings. The CNN architectures like MobileNetV2, ResNet101V2, and DenseNet169 have unparalleled ability to capture intricate spatial relationships within images, which is crucial for discerning subtle patterns like challenging signals such as bubbles. This aligns with the evolving field of diagnostics, which is moving towards inventing and incorporating more versatile readouts like bubbles to allow for more sensitive and unique detection capabilities, distinct from common ones like color and fluorescence. These CNN architectures offer distinct trade-offs in terms of model size, computational efficiency, and classification accuracy, offering flexibility in addressing the specific nuances of the dataset.

To investigate the capabilities of the selected set of ML and DL algorithms in testing microfluidics, we captured 19,097 images of our microfluidic model with bubbles in various settings, including different environments, lighting conditions, times of the day, and backgrounds (**Figure 1**). We labeled the captured images either positive or negative, based on the number of bubbles, around a threshold value of 10 bubbles per microchip, to train our ML and DL models (**Figure 1a**). Out of the 19,097 labelled images (**Figure 1b**), 15,530 images were utilized for training using Python running on Lambda Vector GPU Workstation (Intel i9-10900x CPU, NVIDIA RTX A6000 GPU) system.

**Figure 1.**
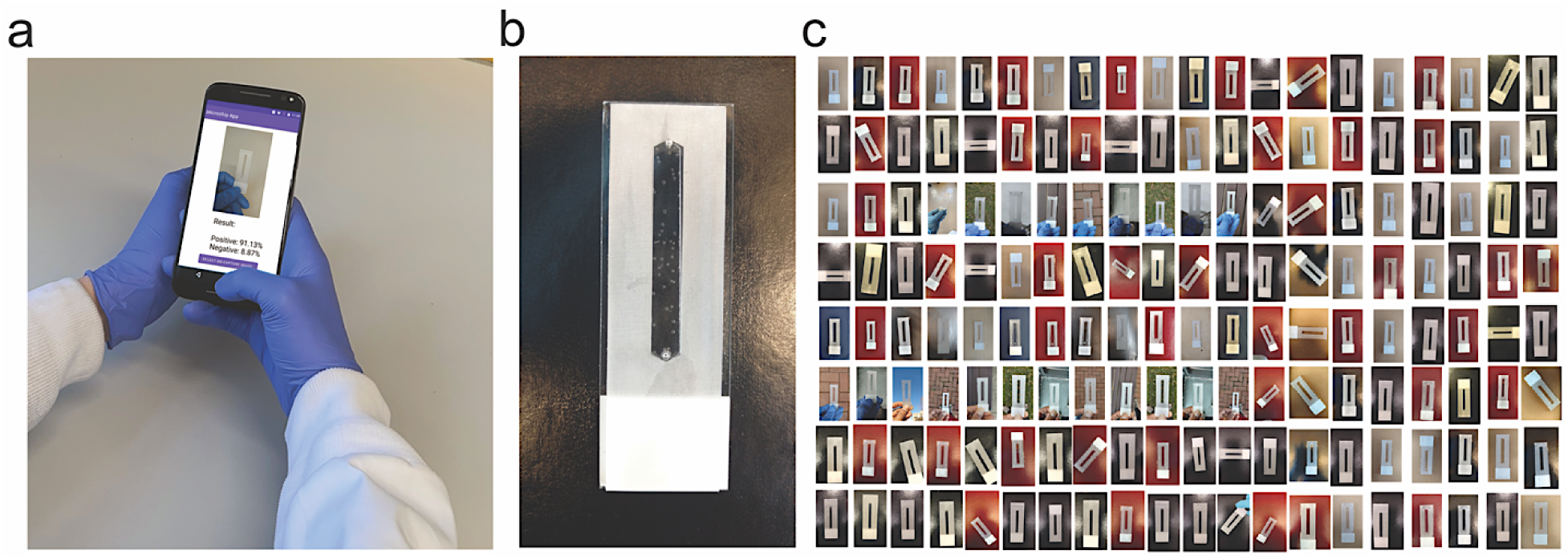
AI algorithms integration and the tested microfluidic model system. (**a**) Microfluidics testing using an integrated POC compatible system running AI algorithm on a cellphone. The system supports a broad range of AI algorithms including both machine learning (ML) and deep learning (DL) models. (**b**) The developed microfluidic model with a single microfluidic channel (length 42 mm, width 5 mm and height 100 µm) containing platinum nanoparticle-seeded bubbles of variable shapes and sizes. (**c**) Snapshot of the image library of the tested microfluidic model collected using cellphone POC system (161 randomly selected images out of 19,097), illustrating the diversity of color, background and brightness.

To test the performance of ML models, we used 1595 randomly selected images, excluding those used for training, to evaluate their classification accuracy. We employed standard performance metrics, including accuracy, precision, recall (i.e., sensitivity), specificity, F1 score, and Matthews’s correlation coefficient (MCC) (**Supplementary Table 1**), obtained from each model to determine their effectiveness[51]. We conducted all statistical analyses and data visualizations using TensorFlow and TensorBoard tools with necessary Python libraries as matplotlib, NumPy, Keras, Sklearn, pandas, torch[52, 53]. The comparison primarily centered around specificity and sensitivity values, which are metrics influencing overall performance and gives information about other metrics.

Our analysis of the ML models revealed that logistic regression and Random Forest models exhibited exceptional sensitivity (>90%), while K-nearest neighbors and Random Forest models demonstrated high specificity (>80%) (**Figure 2a**). The results showed that the highest sensitivity value was obtained from the Random Forests (95.52%) and the highest specificity value was obtained from K-nearest neighbors (89.68%) ML models. we assessed the confusion matrix to better understand the positive and negative predictions. Out of 1595 images, 1447 were classified correctly, with 45 false negatives and 103 false positives. The model primarily made errors in the classification of negative samples. (**Figure 2b** and **Supplementary Figure 1**). The ROC analysis of the trained models indicated that the Random Forest (AUC: 97%) (**Figure 2c**) and K-nearest neighbors (AUC: 90%) have highest area under the ROC, which represents the diagnostic ability of the model (**Supplementary Figure 2**). Additionally, the Random Forest model outperformed others in terms of F1 score (92.8%) and accuracy (90.72%). This shows that the Random Forest provides most balanced results between precision and sensitivity with highest accuracy. Consequently, the most effective model was observed as Random Forest with notable metrics as 95.52% sensitivity, 82.57% specificity, 90.72% accuracy, 90.3% precision, 92.8% F1 score, 79.95% MCC, and 97% AUC (**Supplementary Table 1**).

**Figure 2.**
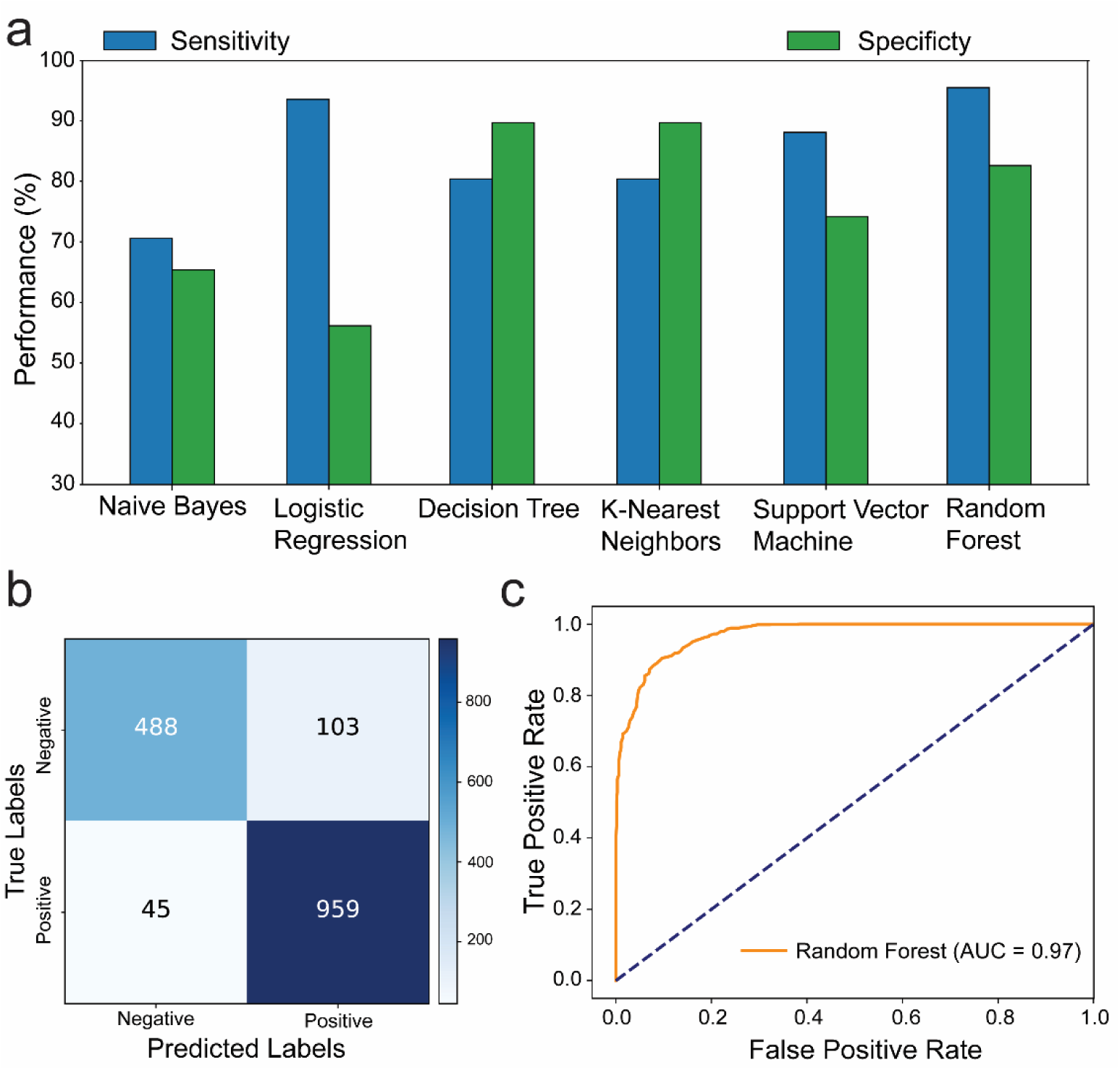
Performance evaluation of machine learning in testing microfluidics. (**a**) Barplots showing the performance (sensitivity and specificity) of the tested ML algorithms (n = 6). All algorithms were trained on our dataset of 15,530 images to classify the model microfluidic chip system with bubble signal into positive or negative around the threshold value of 10 bubbles. (**b**) Confusion matrix showing the number of true negative, false positive, false negative and true positive results when comparing the interpretation of Random Forest ML algorithm to the ground truth classification results. (**c**) ROC analysis of Random Forest performance in testing the model microfluidic chip with bubble signal.

To test the performance of DL models, we continued by evaluating the performance of the selected CNNs architectures using the same dataset of 1595 images. The performance evaluation step was conducted using developed Python algorithms with the help of Pandas, NumPy, Sklearn, Matplotlib, Keras and Tensorflow libraries[52]. The deep learning models utilized for this evaluation included MobileNetV2, EfficientNetV2B0, EfficientNetV2B2, DenseNet169, DenseNet201, InceptionV3, ResNet50V2, EfficientNetB5, and ResNet101V2. In selecting these deep learning models, we prioritized those that does not require significant computing power and thus ensure compatibility for evaluation and testing microfluidics at POC. We also ensured that the chosen models were commonly employed for computer vision tasks, prioritizing ease of integration and robust performance on POC compatible mobile devices[22].

Our results indicated that DenseNet169, EfficientNetB5, and EfficientNetV2B0 exhibited outstanding sensitivity values of 92.63%, 95.82%, and 91.93%, respectively (**Figure 3a and Supplementary Figure 3-5**). ResNet50V2 (89.17%) and InceptionV3 (88.49%) demonstrated high specificity values, while DenseNet169 displayed an exceptional specificity of 92.22% (**Supplementary Table 2**). The confusion matrix revealed further insights into the performance of these algorithms. DenseNet169 algorithm excelled in detecting negative samples, accurately classifying 545 out of 591, while also achieving the second-highest performance in positive classification with 930 out of 1004, resulting in the highest overall performance at 92% (**Figure 3b**). Other algorithms including EfficientNetB5 correctly identified 962 out of the tested 1004 positive samples. However, it misclassified 293 negative samples as positive, resulting in a 50.4% performance rate for negative samples and an overall performance rate of 79%. EfficientNetV2B0 exhibited similar performance, albeit with a 7% overall performance rate downgrade, reflecting a 4% difference in true positive performance rate and an 11% decrease in true negative performance rate. The results of MobileNetV2, EfficientNetV2B2, DenseNet201, InceptionV3, ResNet50V2, and ResNet101V2 algorithms are shown in **Supplementary Figure 4, 5** with misclassification rates < 38%. The ROC analysis of the trained DL models, ResNet50V2 (AUC: 96%), ResNet101V2 (AUC: 96%), InceptionV3 (AUC: 95%) and DenseNet169 (AUC: 92%) and DenseNet201 (AUC: 90%) had the highest area under the ROC (**Supplementary Figure 6, 7**). Additionally, the DenseNet169 model outperformed other models in terms of F1 score (93.94%) and accuracy (92.48%) (**Supplementary Table 2**). Overall, DenseNet169 outperformed other models with the performance metrics and gives the applicable model with 0.92 AUC (**Figure 3c**).

**Figure 3.**
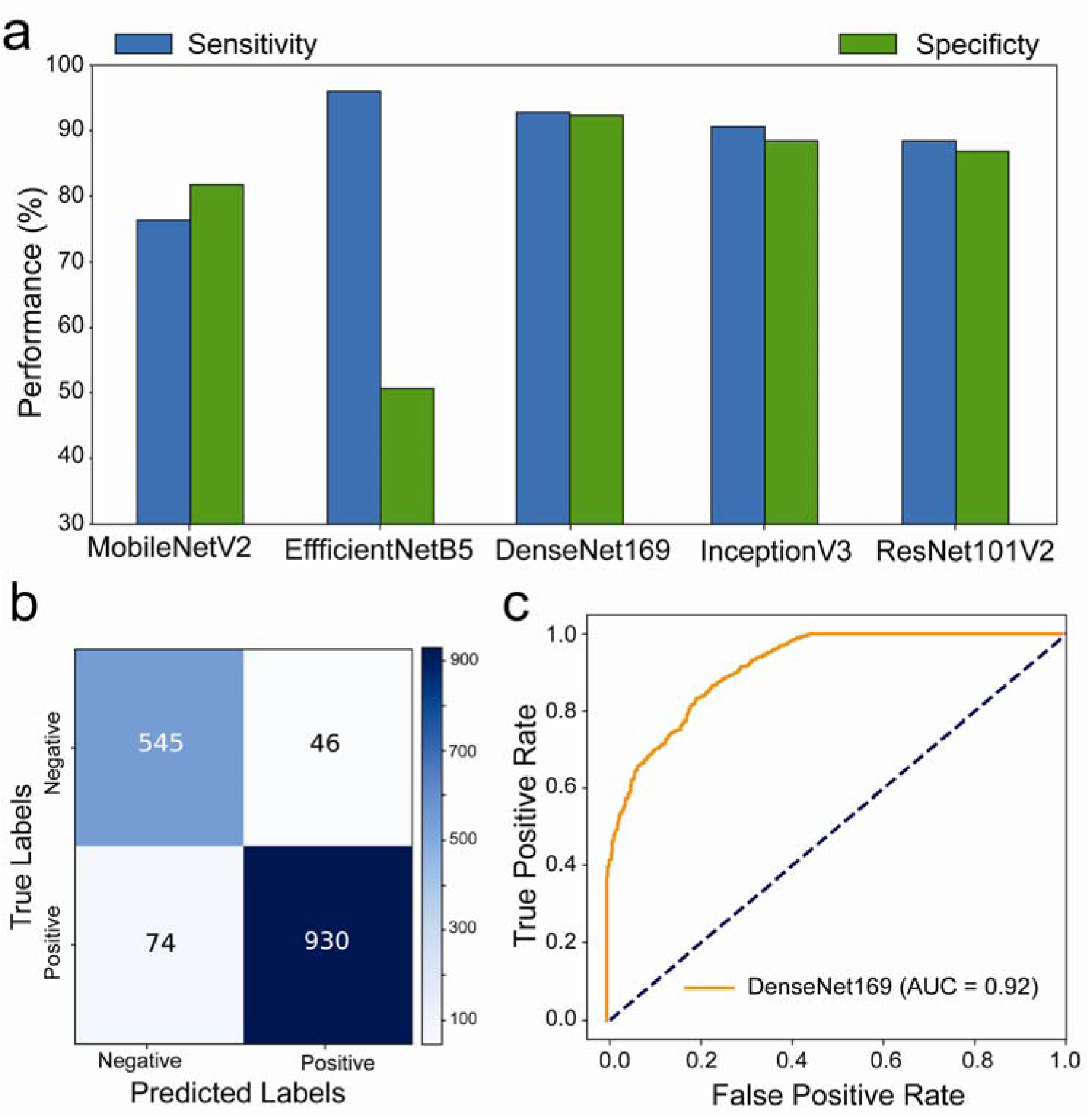
Performance evaluation of deep learning in testing microfluidics. (**a**) Barplots showing the performance (sensitivity and specificity) of the tested DL algorithms (n = 5). All algorithms were trained on our dataset of 15,530 images to classify the model microfluidic chip system with bubble signal into positive or negative around the threshold value of 10 bubbles. (**b**) Confusion matrix showing the number of true negative, false positive, false negative and true positive results when comparing the interpretation of DenseNet169 DL algorithm to the ground truth classification results. (**c**) ROC analysis of DenseNet169 performance in testing the model microfluidic chip system with bubble signal.

We compared the performance of Random Forest and DenseNet169, as these models had outperformed others in our evaluations. To challenge them further, we used a set of 184 microchips prepared with varying numbers of bubbles. A new test set of images was created under different environmental conditions than those used during training. This test set included images taken against different backgrounds (including black, red, brown, metallic grey, and dark blue), rotation, and brightness. This approach allowed us to assess user experience in suboptimal conditions, ensuring a thorough and comprehensive evaluation of the models’ performance in real-world microchip testing scenarios. The generated positive and negative prediction rates were analyzed against the ground truth values of bubbles per chip to evaluate the performance of each model. The results revealed that the DenseNet169 DL model achieves prediction rates with better performance compared to the Random Forest ML model with 80.4% and 88.2% accuracy; 77.98% and 91.81% precision; 81.51% and 87.84% F1 score; 75.3% and 92.31% specificity; and 61.03% and 76.69% MCC for Random Forest and DenseNet169, respectively. The confusion matrix and ROC analyses, on the other hand, confirmed that the DenseNet169 DL algorithm is the optimal prediction model for testing our microfluidic model, outperforming the Random Forest ML algorithm by 87% in AUC and 92% in accuracy classifying true positive and true negative (**Figure 4b, c**).

**Figure 4.**
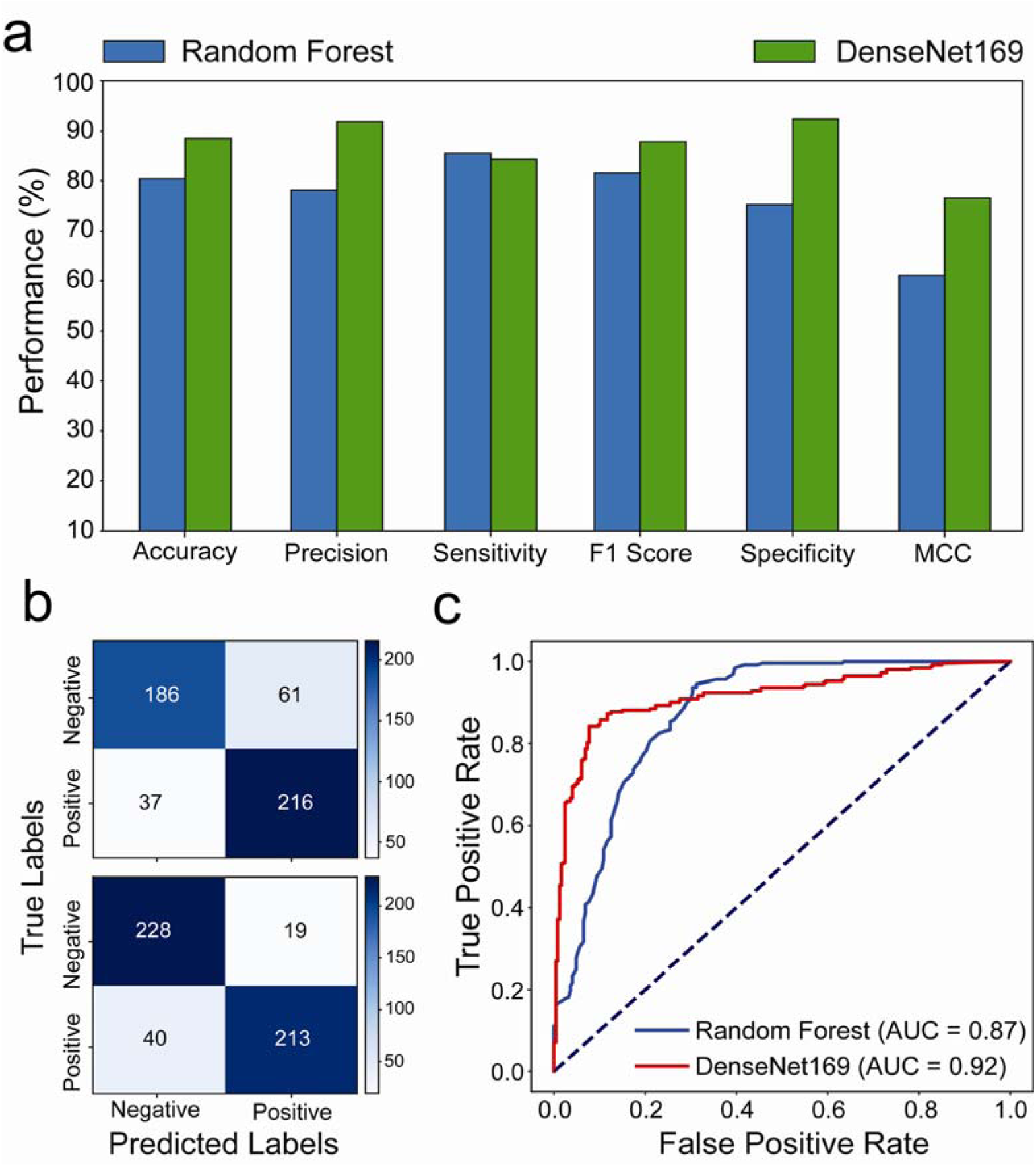
Performance evaluation of machine learning compared to deep learning in testing microfluidics under POC settings. (**a**) Performance matrices (accuracy, precision, sensitivity, F1 score, specificity, and MCC) of the Random Forest ML and the DenseNet169 DL in testing the model microfluidic chip system under challenging imaging conditions that simulate POC testing settings (i.e., different backgrounds, brightness, resolution, cameras, and rotations). (**b**) Confusion matrices showing the number of true negative, false positive, false negative and true positive results when comparing the interpretation of the Random Forest ML and the DenseNet169 DL algorithms to the ground truth classification results. (**c**) ROC analysis of the Random Forest ML and the DenseNet169 DL algorithms performance in testing the model microfluidic chip system with bubble signal.

To demonstrate the effectiveness of incorporating AI in real-world sample testing scenarios using POC-compatible systems, a mobile application capable of running the DenseNet169 model seamlessly was developed, without the need for further optimization. The application features a simple interface for initiating model evaluation and presents results in terms of positive and negative prediction rates, along with images of the tested microfluidic chips (**Supplementary Figure 8**). Out of 250 images, 212 were classified correctly, 29 were classified as false negatives, and 9 were classified as false positives. The model primarily made errors in classifying positive samples. The performance metrics were as follows: Accuracy: 84.8%, Precision: 93.23%, Sensitivity/Recall: 81.05%, F1 Score: 86.71%, Specificity: 90.72%, and MCC: 70.09. The deep learning model achieved an AUC value of 0.90, highlighting its superiority in testing our microfluidic model with bubbles (**Figure 5b**). Furthermore, upon examining the confusion matrix alongside sensitivity and specificity values. Results showed that the DenseNet169 deep learning model achieved 81.05% sensitivity and 90.72% specificity (**Figure 5a**). Heatmap analysis was conducted using images with bubble counts ranging from 0 to 100. The results indicated a higher margin of error around the threshold of 10 bubbles, particularly chips with around 20 to 30 bubbles are ∼30 % misclassified as negative.

**Figure 5.**
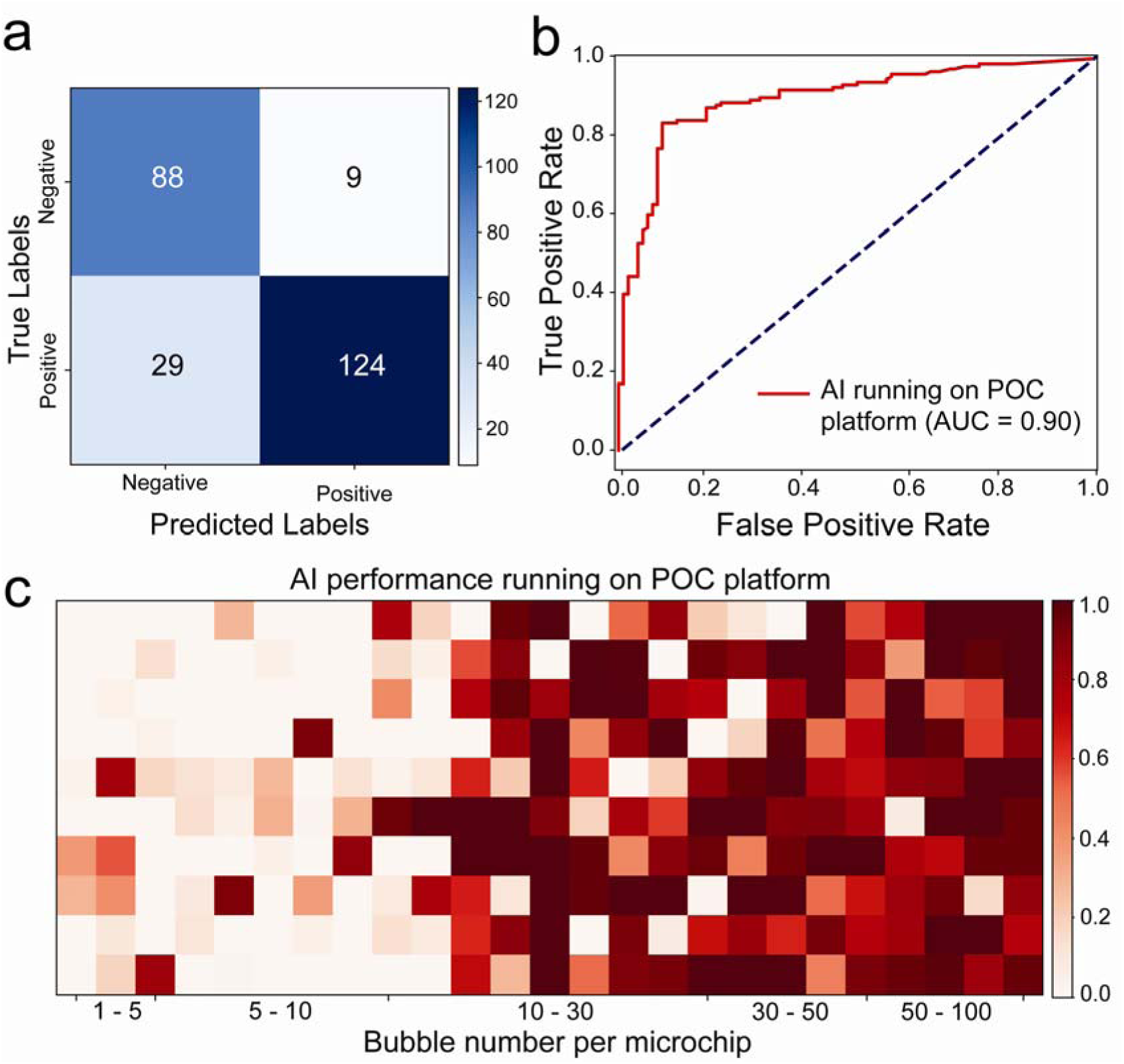
Performance evaluation of AI in testing microfluidics under POC settings using a compatible cellphone system. (**a**) The confusion matrix showing the number of true negative, false positive, false negative and true positive results when comparing AI (i.e., the DenseNet169 DL algorithm) interpretation to the ground truth classification results based on the number of bubbles per microchip. (**b**) ROC analysis of AI performance in testing the model microfluidic chip system with bubble signal. (**c**) Heatmap plot of the probability values of the model microfluidic testing interpretation by AI performance based on the number of bubbles per microchip.

Our study provides a comprehensive evaluation of both ML and deep learning DL algorithms in the context of microfluidics testing under POC settings. Among the ML models, Random Forest emerged as the top performer with a sensitivity of 95.52%, specificity of 82.57%, and an AUC of 97%, showcasing its strong capability in accurately classifying microfluidic device images. The high sensitivity and specificity values underscore Random Forest’s effectiveness in distinguishing positive from negative samples even in challenging imaging conditions. However, the higher rate of false positives indicates a potential area for improvement. In contrast, DL models, particularly DenseNet169, exhibited outstanding performance with sensitivity and specificity values of 92.63% and 92.22%, respectively. DenseNet169’s consistent high performance across different testing conditions, including variations in background and lighting, highlights its robustness and adaptability, making it highly suitable for real-world POC diagnostics where consistent and reliable performance is crucial.

Despite the promising results, several challenges must be addressed to facilitate the widespread adoption of AI in microfluidic POC diagnostics. One key issue is the misclassification of samples with a marginal number of bubbles, especially around the threshold of 10 bubbles, which was evident in the heatmap analysis. Further refinement of the AI models and incorporating additional features or training data will be necessary to enhance accuracy in borderline cases. Combining multiple algorithms can also help overcome these challenges. For example, employing ensemble techniques that integrate models like U-Net for image segmentation and Canny edge detection for edge detection could improve precision in detecting subtle features. Additionally, integrating algorithms such as YOLO (You Only Look Once) for real-time object detection and HOG (Histogram of Oriented Gradients) for robust feature extraction can further enhance the accuracy and reliability of microfluidic POC diagnostics. Such hybrid approaches can leverage the strengths of different algorithms, providing a more comprehensive and accurate analysis.

Moreover, integrating AI models into mobile applications for point-of-care (POC) testing will necessitate ensuring seamless operation across a wide range of devices and environmental conditions, with a strong emphasis on user-friendliness and reliability. This integration is pivotal for achieving the robustness required for practical deployment in diverse healthcare settings. The successful implementation of AI in microfluidic POC diagnostics has far-reaching implications for the healthcare industry, especially in resource-limited settings where access to sophisticated medical infrastructure is often constrained. By enabling rapid, accurate, and on-site testing, AI-driven POC systems address one of the most pressing challenges in modern medicine: the need for timely and precise diagnostics. By democratizing access to high-quality diagnostic tools, AI-integrated POC systems empower frontline healthcare providers with actionable insights, fostering a more equitable distribution of medical resources. This shift supports personalized medicine approaches, tailoring treatment plans to individual patient profiles based on accurate and immediate diagnostic data. Ultimately, the widespread adoption of AI-enhanced microfluidic POC diagnostics can transform healthcare delivery, making it more accessible, efficient, and responsive to the needs of diverse populations worldwide.

### Conclusion

The transformative impact of AI on healthcare is rapidly increasing, particularly in advancing precision medicine through accurate and accessible diagnostics. By conducting a comprehensive comparative evaluation of AI models in testing microfluidics, we have demonstrated the superiority of AI-driven approaches over traditional methods, particularly in the context of POC diagnostics. Through the integration of ML and DL algorithms, we created a diverse ensemble of models capable of leveraging various aspects of the data, thereby enhancing robustness and generalization performance. Our results revealed that the Random Forest ML model and the DenseNet169 DL model exhibited exceptional performance, surpassing other algorithms in terms of sensitivity, specificity, and AUC values. DenseNet169 integration into a mobile POC system demonstrated exceptional accuracy, outperforming traditional visual interpretation by a significant margin. This confirms the potential of AI to revolutionize diagnostics, offering more accurate and efficient testing solutions in resource-limited settings. Moreover, our findings highlight the significant role that AI can play into healthcare systems, as it holds promise for enhancing patient outcomes, streamlining healthcare delivery, and ultimately, democratizing access to high-quality diagnostic services. Moving forward, further research and development efforts are warranted to optimize AI algorithms for real-world deployment, ensuring their seamless integration into clinical practice and maximizing their impact on global health outcomes.

## MATERIAL AND METHODS

### Microfluidic chip model design and fabrication

We developed a microfluid chip system that features a single microfluidic channel. The microchip was designed using the vector graphics editor CorelDRAW Graphics suite software, and fabricated of polymethyl methacrylate (PMMA) (3.125 mm thick), DSA film (100 μm thick, 3M, USA), and glass slides (25 mm x 75 mm). The fabrication process starts by cutting PMMA and DSA film using a laser cutter (Boss Laser LS-1416, USA). The PMMA was prepared to contain the microfluidic channel inlet and outlet, while DSA film included the main testing channel. All materials were precleaned with 70% ethanol, and deionized water using lint-free tissue. The surface of the cleaned glass slides was treated and cleaned using oxygen plasma (PE-25, 100 mW, 15% oxygen; Plasma Etch Inc.) for 10 minutes. Then PMMA and DSA film were assembled on the modified glass slide, forming the model microfluidic chip system. Each system was loaded with platinum nanoparticle-seeded bubbles. PtNPs synthesized using our previously published protocol were mixed with a peroxide-containing solution (5% hydrogen peroxide and 20% glycerol) and loaded on chip system. The concentration of added PtNPs was controlled to prepare systems with variable numbers of bubbles (0 – >200 bubbles per chip), randomly distributed within the microfluidic channel.

### AI models selection, training and performance testing

We selected a set of 15 models that encompass a number of machine learning and deep learning models, widely reported to have high performance in image classification and pattern recognition. The machine learning models included Naive Bayes, Logistic Regression, Decision Tree, K-Nearest Neighbors, Support Vector Machine and Random Forest, while the deep learning models of MobileNetV2, EfficientNetV2B0, EfficientNetV2B2, DenseNet169, DenseNet201, InceptionV3, ResNet50V2, EfficientNetB5 and ResNet101V2, were selected to support workflow running on mobile devices and systems. We generated a dataset of 19,097 images of the model microfluidic system captured using Moto XT1575, iPhone X and Vivo smartphones. The dataset comprises two groups, i.e., positive (with > 10 bubbles per microchip) and negative (in range of < 10 bubbles per microchip) sample images. The microfluidic system imaging was performed at different angles (0 – 360°) and backgrounds and environments to maximize the variations, and make our dataset more robust and comprehensive. We used 15530 images for training, 1788 images for validation and 1012 images for testing the performance of the selected ML and DL models in testing the model microfluidic system and classifying samples into positive and negative based on bubble signal. We started the process by importing pre-trained models available from Scikit-learn and Keras libraries to develop the selected ML and DL models, respectively. In the pre-processing step, the images of our training dataset were resized to the input dimensions of the selected models, leveraging the features learned by ImageNet pretrained network. We performed the batch normalization then used Adam optimizer to fine-tune the network using a global learning rate of 0.001. In addition, we employed a varied number of epochs to test the algorithms optimal performance and we set the number to 50 epochs. Then we performed the transfer learning by removing the final classification layer from the chosen networks and trained with our dataset. All the algorithms were trained on Vector Workstation (Intel i9-10900x CPU and NVIDIA RTX A6000 GPU, Lambda) and after training, we tested the performance of the best-performing ML and DL algorithms individually using a challenging dataset of 400 images. This testing dataset included rotated images, images with various colored backgrounds (matte, bright, reflective), and images with lens distortion and brightness variations. The ML algorithms were evaluated using the sklearn and torch libraries, while the DL algorithm was evaluated using the TensorFlow library. Performance metrics such as accuracy, precision, sensitivity, and F1-score were employed to quantitatively measure classification accuracy and the ability of each model to correctly identify the tested microchip.

### AI testing on a POC compatible system

We utilized the open-source platform Android Studio (version Giraffe 2022.3.1) to develop an AI-enabled mobile application. Android Studio offers an integrated development environment (IDE) tailored for Android application development. The application facilitates the capture of sensor images through the smartphone’s built-in camera or from images stored in the device’s memory. A trained DL model, DenseNet169, was converted to TensorFlow Lite and integrated into the application, which was developed for Android 6.0 (API level 23). This application was installed on a Moto XT1575 and used as a proof-of-concept system for testing microfluidics with images simulating real-world conditions. We evaluated the performance of the AI model using a testing set of 250 images, each featuring 0-100 bubbles per chip. This testing set included images with challenging backgrounds and imaging conditions, such as noise, blur, hand interaction, daylight, artificial light, natural and artificial occlusion, resolution variability, and the presence of small bubbles. The classification results, displayed on the user interface, indicate the probability of a sample being positive (>50%) or negative (<50%). The correlation between AI-generated classification results and the number of bubbles per chip was analyzed, and prediction accuracy rates were employed to generate performance metrics.

## Data Availability

All data produced in the present work are contained in the manuscript

## References

1. Rajpurkar, P., et al., AI in health and medicine. Nature Medicine, 2022. 28(1): p. 31–38.

2. Esteva, A., et al., A guide to deep learning in healthcare. Nature medicine, 2019. 25(1): p. 24–29.

3. Topol, E.J., High-performance medicine: the convergence of human and artificial intelligence. Nature medicine, 2019. 25(1): p. 44–56.

4. Acosta, J.N., et al., Multimodal biomedical AI. Nature Medicine, 2022. 28(9): p. 1773–1784.

5. Hosny, A., et al., Artificial intelligence in radiology. Nature Reviews Cancer, 2018. 18(8): p. 500–510.

6. Kermany, D.S., et al., Identifying medical diagnoses and treatable diseases by image-based deep learning. cell, 2018. 172(5): p. 1122–1131. e9.

7. Aggarwal, R., et al., Diagnostic accuracy of deep learning in medical imaging: a systematic review and meta-analysis. npj Digital Medicine, 2021. 4(1): p. 65.

8. Lambin, P., et al., Radiomics: the bridge between medical imaging and personalized medicine. Nature Reviews Clinical Oncology, 2017. 14(12): p. 749–762.

9. Sermesant, M., et al., Applications of artificial intelligence in cardiovascular imaging. Nature Reviews Cardiology, 2021. 18(8): p. 600–609.

10. Oren, O., B.J. Gersh, and D.L. Bhatt, Artificial intelligence in medical imaging: switching from radiographic pathological data to clinically meaningful endpoints. The Lancet Digital Health, 2020. 2(9): p. e486–e488.

11. Yager, P., G.J. Domingo, and J. Gerdes, Point-of-care diagnostics for global health. Annu. Rev. Biomed. Eng., 2008. 10: p. 107–144.

12. Chan, C.P.Y., et al., Evidence-based point-of-care diagnostics: current status and emerging technologies. Annual review of analytical chemistry, 2013. 6: p. 191–211.

13. Wang, C., et al., Point-of-care diagnostics for infectious diseases: From methods to devices. Nano Today, 2021. 37: p. 101092.

14. Riordon, J., et al., Deep learning with microfluidics for biotechnology. Trends in biotechnology, 2019. 37(3): p. 310–324.

15. Chen, S., et al., Wearable flexible microfluidic sensing technologies. Nature Reviews Bioengineering, 2023. 1(12): p. 950–971.

16. Zhou, J., et al., High-throughput microfluidic systems accelerated by artificial intelligence for biomedical applications. Lab on a Chip, 2024.

17. Zhao, W., et al., Computer vision-based artificial intelligence-mediated encoding-decoding for multiplexed microfluidic digital immunoassay. ACS nano, 2023. 17(14): p. 13700–13714.

18. Ao, Z., et al., Microfluidics guided by deep learning for cancer immunotherapy screening. Proceedings of the National Academy of Sciences, 2022. 119(46): p. e2214569119.

19. Draz, M.S. and H. Shafiee, System and method for virus detection using nanoparticles and a neural network enabled mobile device. 2023, Google Patents.

20. Draz, M.S., et al., DNA engineered micromotors powered by metal nanoparticles for motion based cellphone diagnostics. Nature communications, 2018. 9(1): p. 1–13.

21. Draz, M.S., et al., Motion-based immunological detection of Zika virus using Pt-nanomotors and a cellphone. ACS nano, 2018. 12(6): p. 5709–5718.

22. Wang, B., et al., Smartphone-based platforms implementing microfluidic detection with image-based artificial intelligence. Nature Communications, 2023. 14(1): p. 1–18.

23. Liu, H., et al., Functions and applications of artificial intelligence in droplet microfluidics. Lab on a Chip, 2023. 23(11): p. 2497–2513.

24. Zheng, J., et al., Exploiting machine learning for bestowing intelligence to microfluidics. Biosensors and Bioelectronics, 2021. 194: p. 113666.

25. Bi, W.L., et al., Artificial intelligence in cancer imaging: clinical challenges and applications. CA: a cancer journal for clinicians, 2019. 69(2): p. 127–157.

26. Arora, N., A.K. Banerjee, and M.L. Narasu, The role of artificial intelligence in tackling COVID-19. 2020, Future Medicine. p. 717–724.

27. Smith, K.P., et al., Applications of artificial intelligence in clinical microbiology diagnostic testing. Clinical Microbiology Newsletter, 2020. 42(8): p. 61–70.

28. Smith, K.P. and J.E. Kirby, Image analysis and artificial intelligence in infectious disease diagnostics. Clinical Microbiology and Infection, 2020. 26(10): p. 1318–1323.

29. London, A.J., Artificial intelligence and black-box medical decisions: accuracy versus explainability. Hastings Center Report, 2019. 49(1): p. 15–21.

30. Gilvary, C., et al., The missing pieces of artificial intelligence in medicine. Trends in pharmacological sciences, 2019. 40(8): p. 555–564.

31. Dayhoff, J.E. and J.M. DeLeo, Artificial neural networks: opening the black box. Cancer: Interdisciplinary International Journal of the American Cancer Society, 2001. 91(S8): p. 1615–1635.

32. Koteluk, O., et al., How do machines learn? artificial intelligence as a new era in medicine. Journal of Personalized Medicine, 2021. 11(1): p. 32.

33. Bhaskar, H., D.C. Hoyle, and S. Singh, Machine learning in bioinformatics: A brief survey and recommendations for practitioners. Computers in biology and medicine, 2006. 36(10): p. 1104–1125.

34. de Hond, A.A., et al., Guidelines and quality criteria for artificial intelligence-based prediction models in healthcare: a scoping review. NPJ digital medicine, 2022. 5(1): p. 2.

35. Zhuang, F., et al., A comprehensive survey on transfer learning. Proceedings of the IEEE, 2020. 109(1): p. 43–76.

36. Pan, S.J. and Q. Yang, A survey on transfer learning. IEEE Transactions on knowledge and data engineering, 2009. 22(10): p. 1345–1359.

37. Ooge, J., G. Stiglic, and K. Verbert, Explaining artificial intelligence with visual analytics in healthcare. Wiley Interdisciplinary Reviews: Data Mining and Knowledge Discovery, 2022. 12(1): p. e1427.

38. Moulaei, K., et al., Generative artificial intelligence in healthcare: A scoping review on benefits, challenges and applications. International Journal of Medical Informatics, 2024: p. 105474.

39. Deo, R.C., Machine learning in medicine. Circulation, 2015. 132(20): p. 1920–1930.

40. Alanazi, A., Using machine learning for healthcare challenges and opportunities. Informatics in Medicine Unlocked, 2022. 30: p. 100924.

41. Morid, M.A., A. Borjali, and G. Del Fiol, A scoping review of transfer learning research on medical image analysis using ImageNet. Computers in biology and medicine, 2021. 128: p. 104115.

42. Salehi, A.W., et al., A study of CNN and transfer learning in medical imaging: Advantages, challenges, future scope. Sustainability, 2023. 15(7): p. 5930.

43. Rahaman, M.M., et al., Identification of COVID-19 samples from chest X-Ray images using deep learning: A comparison of transfer learning approaches. Journal of X-ray Science and Technology, 2020. 28(5): p. 821–839.

44. Tang, X., The role of artificial intelligence in medical imaging research. BJR| Open, 2019. 2(1): p. 20190031.

45. Hasani, N., et al., Trustworthy artificial intelligence in medical imaging. PET clinics, 2022. 17(1): p. 1–12.

46. Angehrn, Z., et al., Artificial intelligence and machine learning applied at the point of care. Frontiers in pharmacology, 2020. 11: p. 759.

47. Yang, Y., et al., Artificial intelligence-assisted smartphone-based sensing for bioanalytical applications: A review. Biosensors and Bioelectronics, 2023: p. 115233.

48. Xu, D., et al., Automatic smartphone-based microfluidic biosensor system at the point of care. Biosensors and Bioelectronics, 2018. 110: p. 78–88.

49. McIntyre, D., et al., Machine learning for microfluidic design and control. Lab on a Chip, 2022. 22(16): p. 2925–2937.

50. Hernández-Neuta, I., et al., Smartphone-based clinical diagnostics: towards democratization of evidence-based health care. Journal of internal medicine, 2019. 285(1): p. 19–39.

51. Jiao, Y. and P. Du, Performance measures in evaluating machine learning based bioinformatics predictors for classifications. Quantitative Biology, 2016. 4: p. 320–330.

52. Stančin, I. and A. Jović. An overview and comparison of free Python libraries for data mining and big data analysis. in 2019 42nd International convention on information and communication technology, electronics and microelectronics (MIPRO). 2019. IEEE.

53. Erickson, B.J., et al., Toolkits and libraries for deep learning. Journal of digital imaging, 2017. 30: p. 400–405.

