## SUPPLEMENTARY for "Artificial Intelligence Performance in Testing Microfluidics for Point-of-Care"

### a. Supplementary Figures.

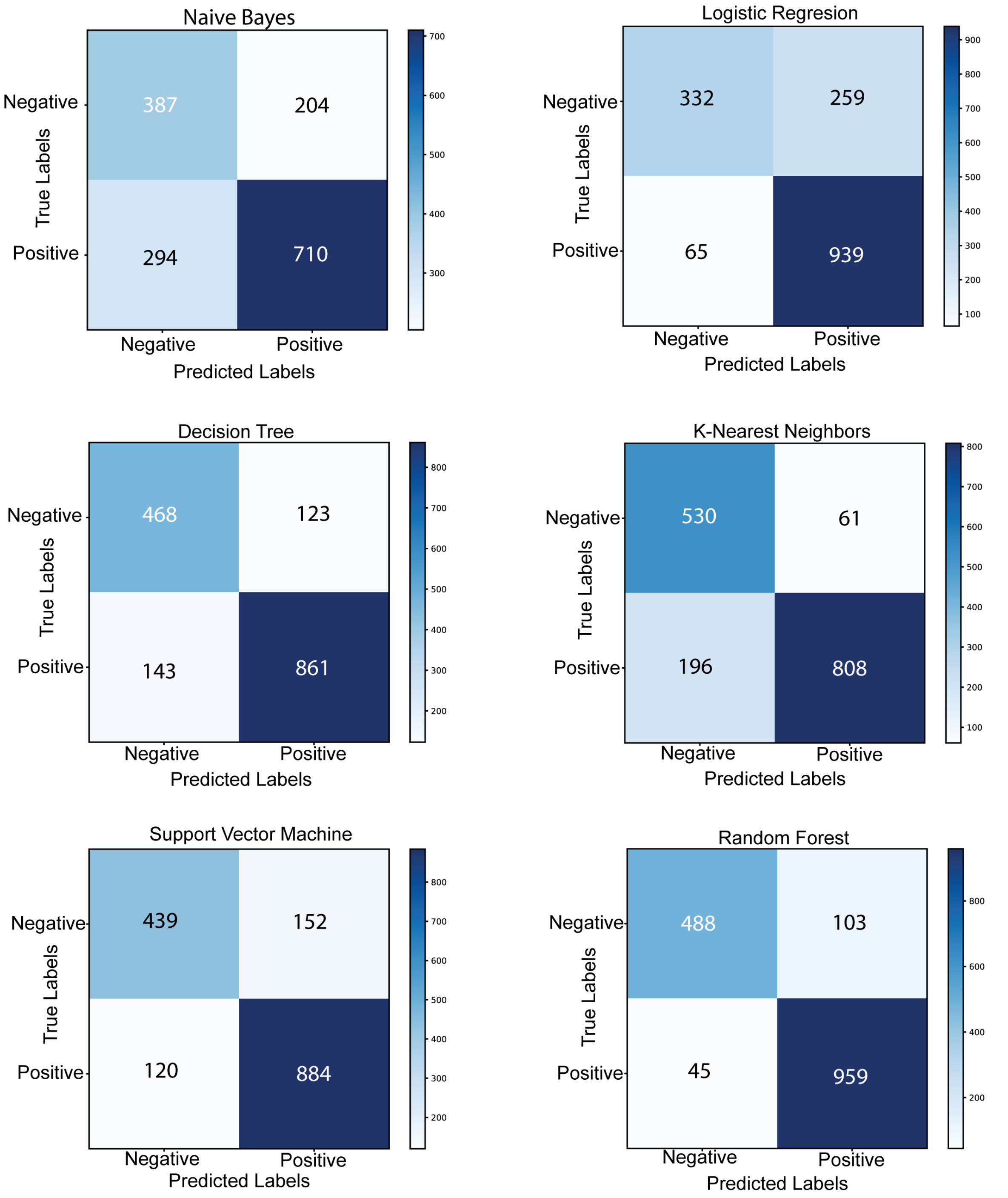


**Supplementary Figure 1.** Confusion matrix analysis of the tested machine learning algorithms.

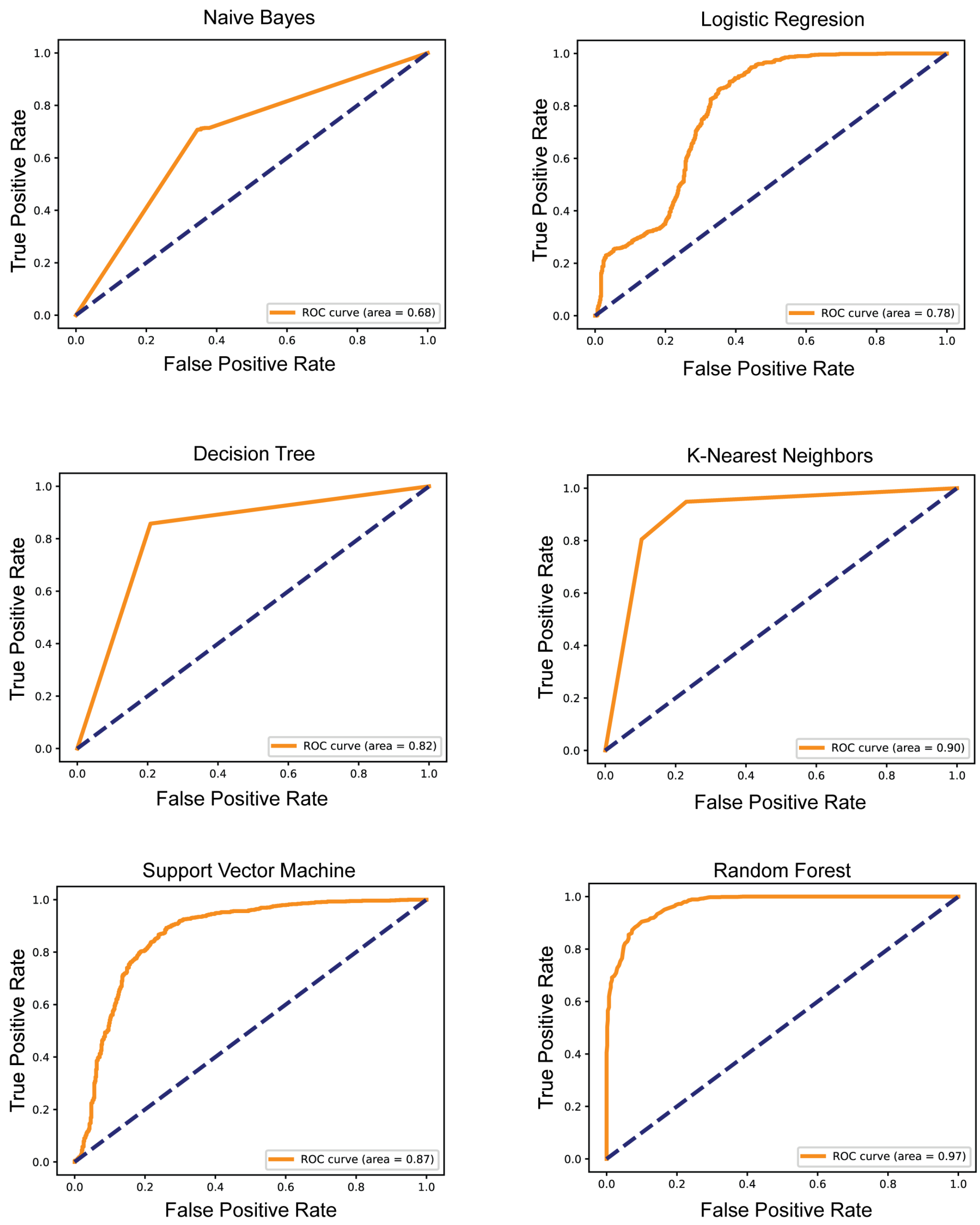


**Supplementary Figure 2**. ROC analysis of the tested machine learning algorithms.

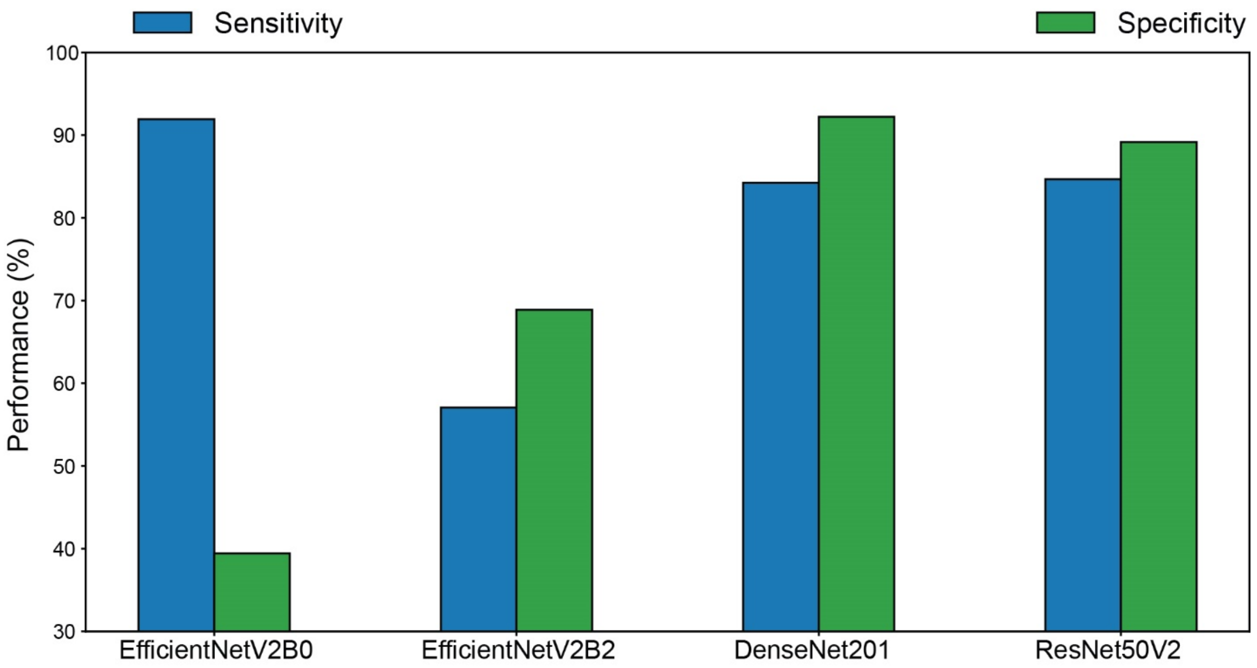


**Supplementary Figure 3**. Performance of deep learning models of EfficientNetV2B0, EfficientNetV2B2, DenseNet201, and ResNet50V2 in microfluidic testing.

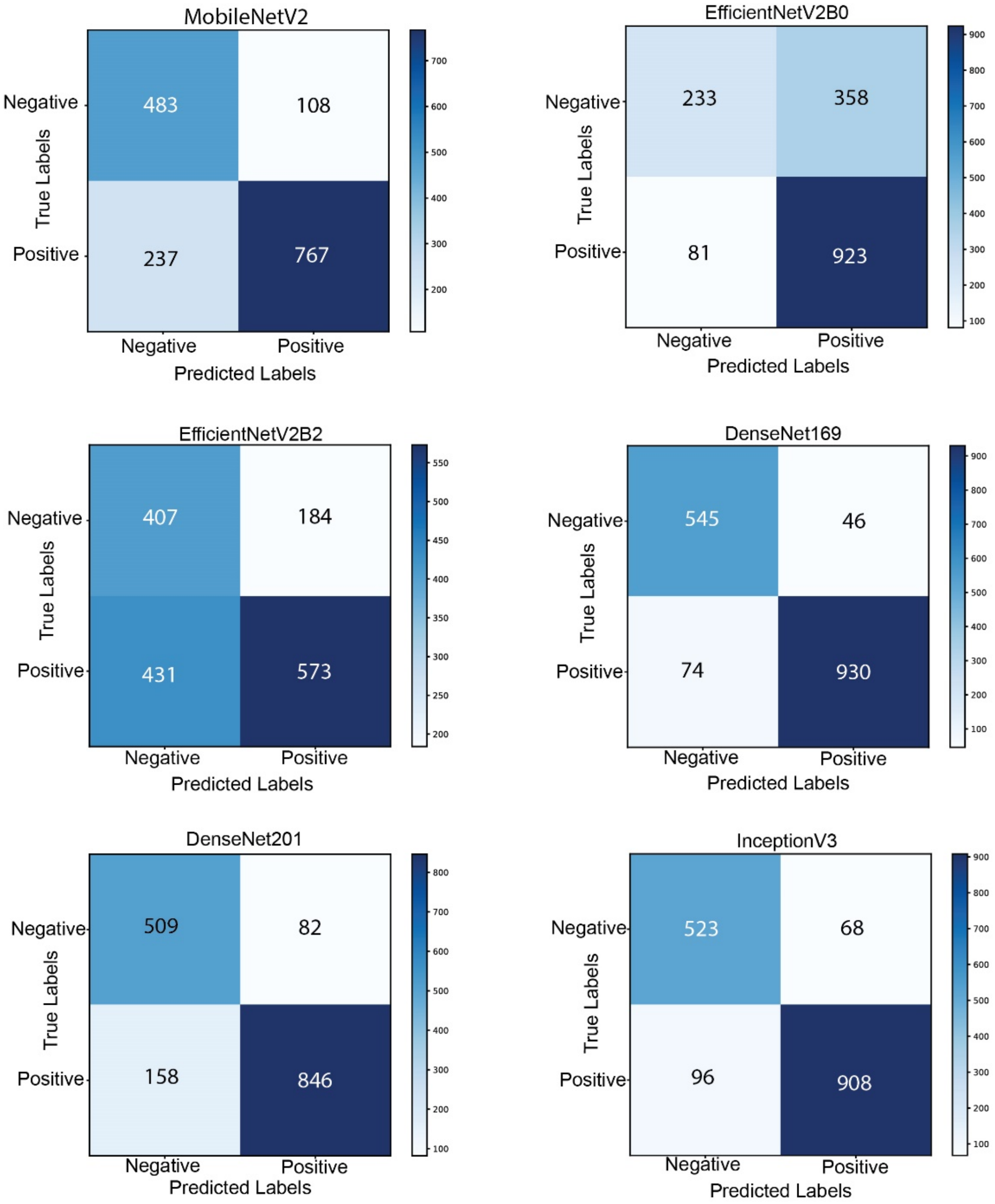


**Supplementary Figure 4.** Confusion matrix analysis of deep learning algorithms of MobileNetV2, EfficientNetV2B0, EfficientNetV2B2, DenseNet169, DenseNet201, and InceptionV3.

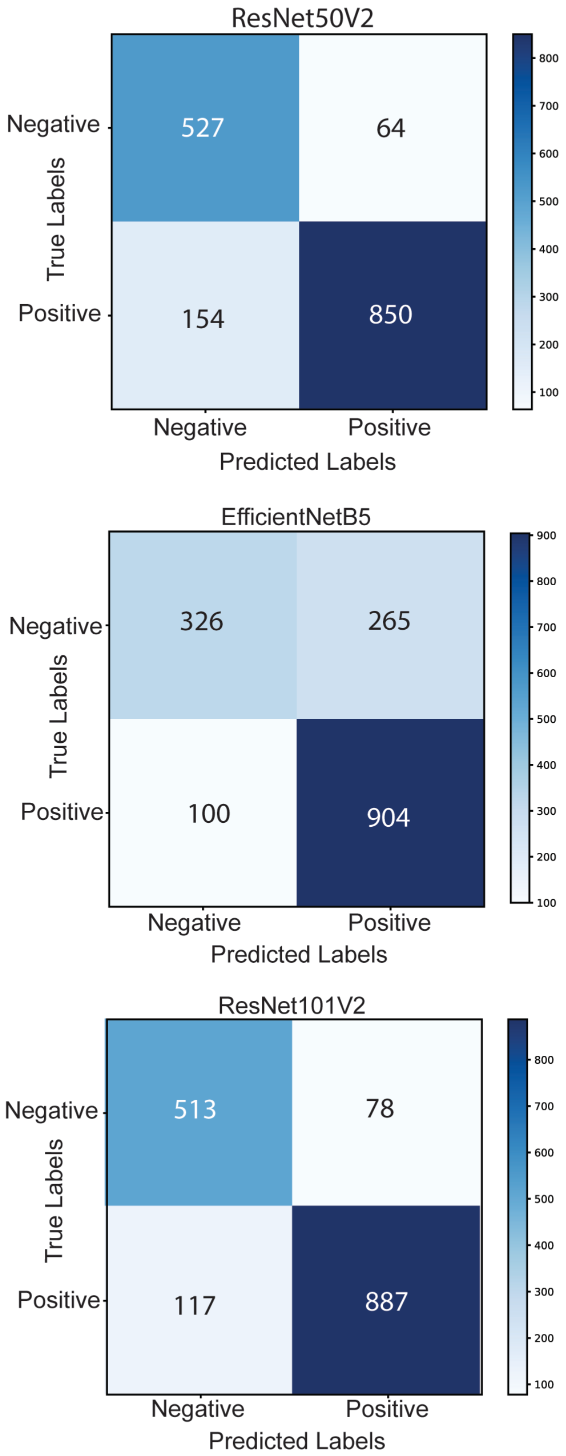


**Supplementary Figure 5.** Confusion matrix analysis of deep learning algorithms of ResNet50V2, EfficientNetB5, and Resnet101V2.

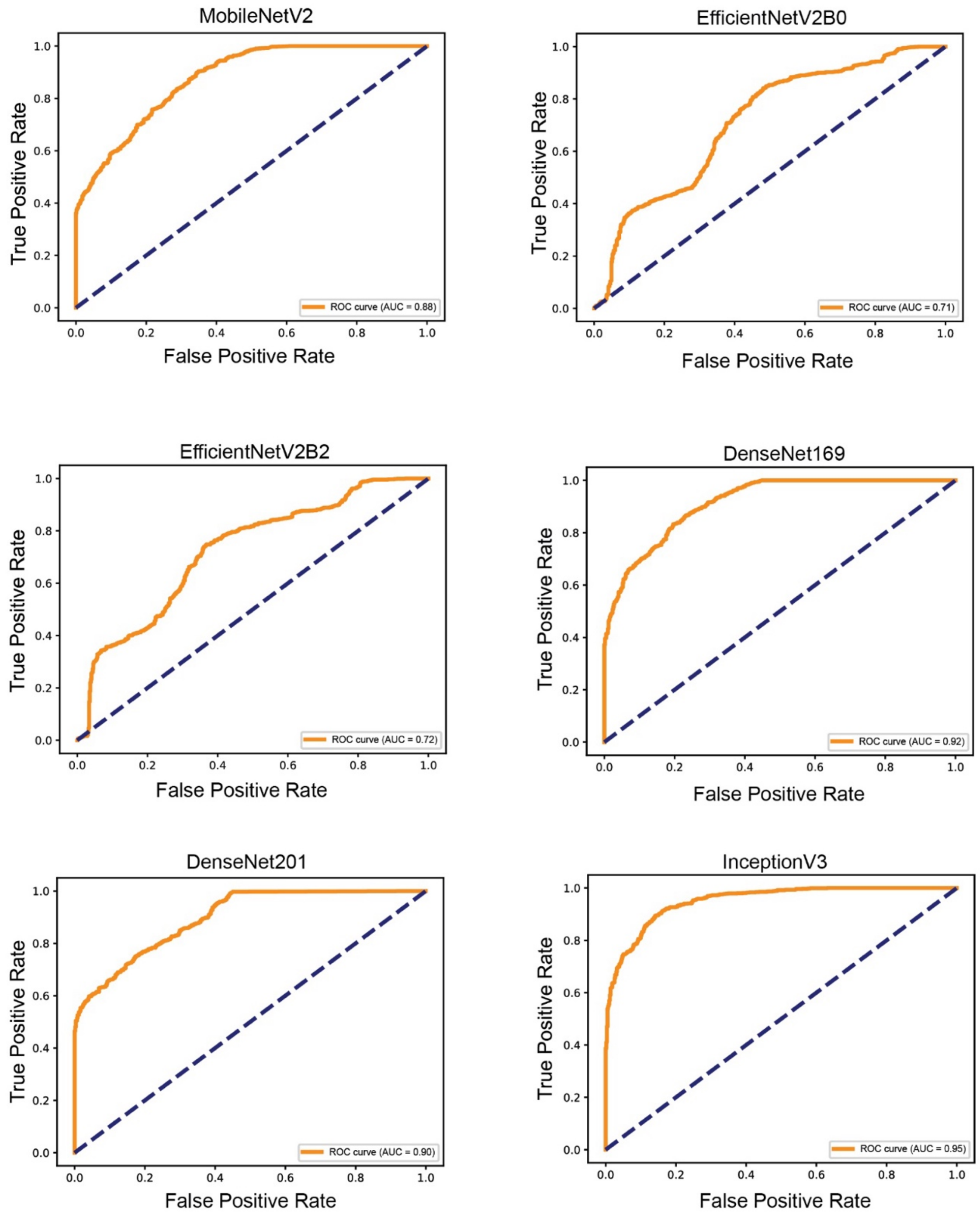


**Supplementary Figure 6.** ROC analysis of deep learning algorithms (MobileNetV2, EfficientNetV2B0, EfficientNetV2B2, DenseNet169, DenseNet201, InceptionV3).

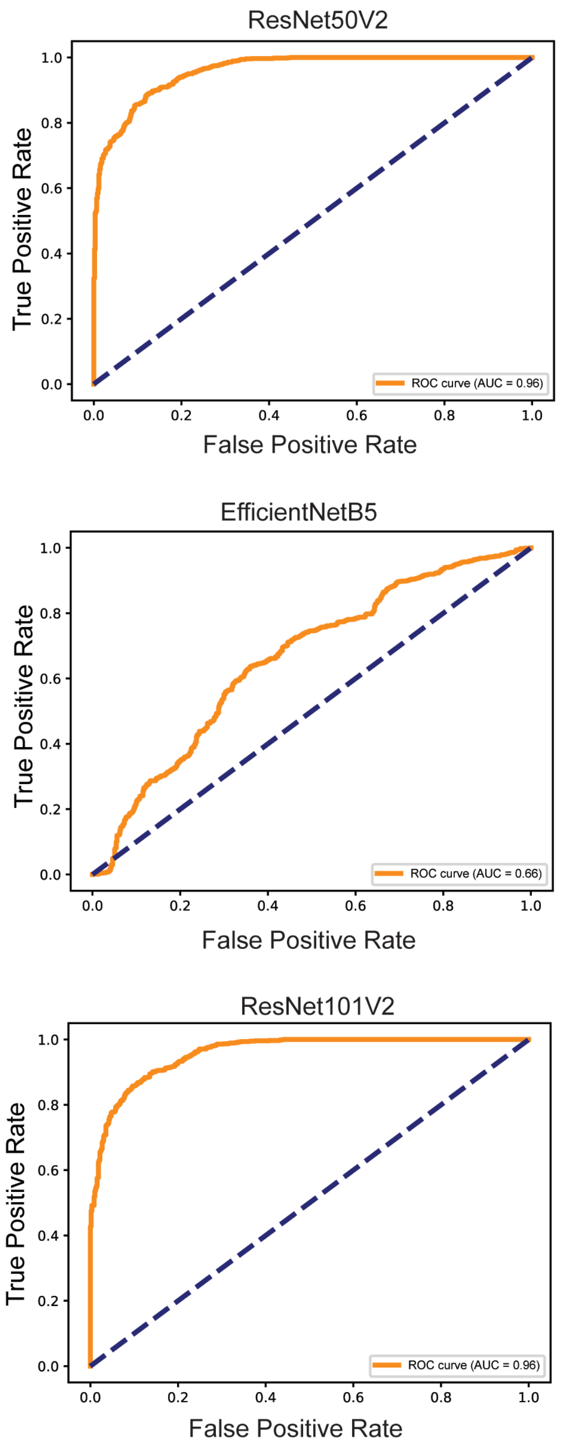


**Supplementary Figure 7.** ROC analysis of deep learning algorithms (ResNet50V2, EfficientNetB5, Resnet101V2).

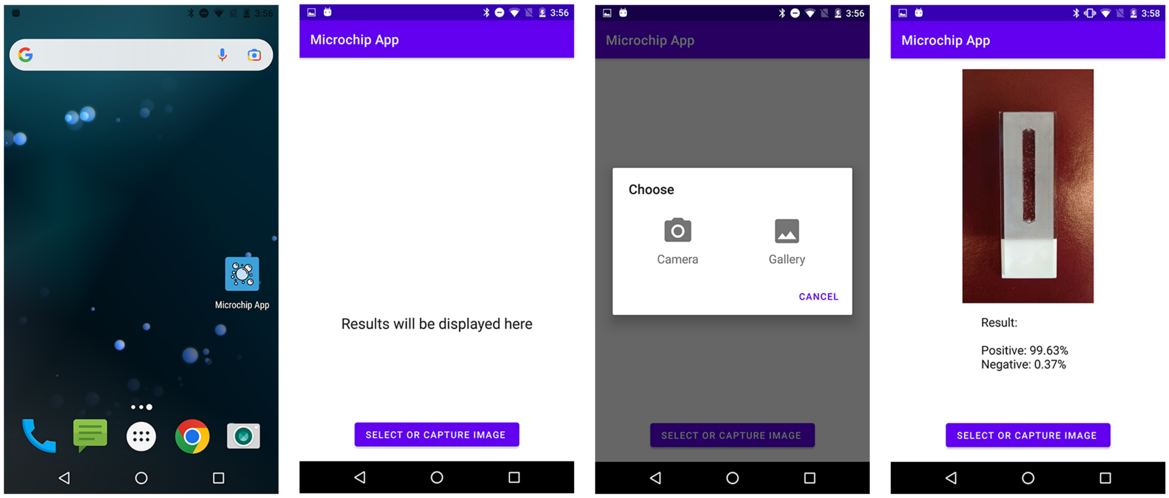


**Supplementary Figure 8.** The mobile application developed with DenseNet169 algorithm, facilitating the testing and classification of microfluidic chips. Users are presented with two distinct options for chip classification: the first option is selecting an image stored on phone, and the second option is starting the cellphone camera and testing sample.

### b. Supplementary Tables

**Supplementary Table 1.** ML algorithms performance in microfluidic testing.

| **Models** | **Accuracy** | **Precision** | **Sensitivity** | **F1 Score** | **Specificity** | **MCC** | **AUC** |
|---|---|---|---|---|---|---|---|
| Naive Bayes | 0.6878 | 0.7768 | 0.7072 | 0.7404 | 0.6548 | 0.3534 | 0.68 |
| Logistic Regression | 0.7969 | 0.7838 | 0.9353 | 0.8529 | 0.5618 | 0.5551 | 0.78 |
| Decision Tree | 0.8332 | 0.875 | 0.8576 | 0.8662 | 0.7919 | 0.6452 | 0.82 |
| K-Nearest Neighbors | 0.8389 | 0.9298 | 0.8048 | 0.8628 | 0.8968 | 0.6804 | 0.9 |
| Support Vector Machine | 0.8295 | 0.8533 | 0.8805 | 0.8667 | 0.7428 | 0.6309 | 0.87 |
| Random Forest | 0.9072 | 0.903 | 0.9552 | 0.928 | 0.8257 | 0.7995 | 0.97 |

**Supplementary Table 2**. DL algorithms performance in microfluidic testing.

| Models | Accuracy | Precision | Sensitivity | F1 Score | Specificity | MCC | AUC |
|---|---|---|---|---|---|---|---|
| MobileNetV2 | 0.7837 | 0.8766 | 0.7639 | 0.8164 | 0.8173 | 0.5641 | 0.88 |
| EfficienNetV2B0 | 0.7248 | 0.7205 | 0.9193 | 0.8079 | 0.3942 | 0.3809 | 0.71 |
| EfficienNetV2B2 | 0.6144 | 0.7569 | 0.5707 | 0.6508 | 0.6887 | 0.2509 | 0.72 |
| DenseNet169 | 0.9248 | 0.9529 | 0.9263 | 0.9394 | 0.9222 | 0.8409 | 0.92 |
| DenseNet201 | 0.8495 | 0.9116 | 0.8426 | 0.8758 | 0.8613 | 0.6892 | 0.9 |
| InceptionV3 | 0.8972 | 0.9303 | 0.9044 | 0.9172 | 0.8849 | 0.7822 | 0.95 |
| ResNet50V2 | 0.8633 | 0.93 | 0.8466 | 0.8863 | 0.8917 | 0.7209 | 0.96 |
| EfficientNetB5 | 0.79 | 0.7665 | 0.9582 | 0.8517 | 0.5042 | 0.5453 | 0.66 |
| ResNet101V2 | 0.8777 | 0.9192 | 0.8835 | 0.901 | 0.868 | 0.7424 | 0.96 |

**Supplementary Table 3**. Performance of ML comparted to DL in microfluidic testing under challenging conditions that simulate real-world sample testing.

| Models | Accuracy | Precision | Sensitivity | F1 Score | Specificity | MCC | AUC |
|---|---|---|---|---|---|---|---|
| **DenseNet169** | 0.882 | 0.9181 | 0.8419 | 0. 8784 | 0. 9231 | 0.7669 | 0.92 |
| **Random Forest** | 0.804 | 0.7798 | 0.8538 | 0.8151 | 0.7530 | 0.6103 | 0.87 |

**Supplementary Table 4.** AI performance in testing microfluidics at POC.

| Models | Accuracy | Precision | Sensitivity | F1 Score | Specificity | MCC | AUC |
|---|---|---|---|---|---|---|---|
| **App** | 0.848 | 0.9323 | 0.8105 | 0.8671 | 0.9072 | 0.7009 | 0.90 |
